# Rapid Point-Of-Care Breath Test Predicts Breast Cancer And Abnormal Mammograms in Symptomatic Women

**DOI:** 10.1101/2020.04.07.20042895

**Authors:** Michael Phillips, Therese B Bevers, Linda Hovanessian Larsen, Nadine Pappas, Sonali Pathak

## Abstract

**Background:** Previous studies have reported volatile organic compounds (VOCs) in the breath as biomarkers of breast cancer. These biomarkers may be derived from cancer-associated fibroblasts, in which oxidative stress degrades polyunsaturated fatty acids to volatile alkanes and methylated alkane derivatives that are excreted in the breath. We evaluated a rapid point-of-care test for breath VOC biomarkers as predictors of breast cancer and abnormal mammograms.

**Methods:** We studied 593 women aged ≥ 18 yr referred to three sites for mammography for a symptomatic breast-related concern (e.g. breast mass, nipple discharge). A rapid point-of-care breath testing system collected and concentrated alveolar breath VOCs on a sorbent trap and analyzed them with gas chromatography and surface acoustic wave detection in < 6 min. Breath VOC chromatograms were randomly assigned to a training set or to a validation set. Monte Carlo analysis identified significant breath VOC biomarkers of breast cancer and abnormal mammograms in the training set, and these biomarkers were incorporated into a multivariate algorithm to predict disease in the validation set.

**Results:** Prediction of breast cancer: 50 women had biopsy-proven breast cancer (invasive cancer 41, ductal non-invasive cancer 9) Unsplit data set: Breath VOCs identified breast cancer with 83% accuracy (area under curve of receiver operating characteristic), 82% sensitivity and 77.1% specificity. Split data sets: Training set breath VOCs identified breast cancer with 80.3% accuracy, 84% sensitivity and 74.3% specificity. Corresponding values in the validation set were 68%% accuracy, 72.4% sensitivity and 61.5% specificity.

Prediction of BIRADS 4 and 5 mammograms (versus BIRADS 1, 2 and 3): Unsplit data set: Breath VOCs identified abnormal mammograms with 76.2% accuracy. Split data sets: Breath VOCs identified abnormal mammograms with 74.2% accuracy, 73.3% sensitivity and 60% specificity. Corresponding values in the validation set were 60.5% accuracy, 64.2% sensitivity and 51% specificity.

**Conclusions:** A rapid point-of-care test for breath VOC biomarkers accurately predicted risk of breast cancer and abnormal mammograms in women with breast-related symptoms.

## INTRODUCTION

Breath contains volatile organic compounds (VOCs) that are biomarkers of breast cancer^1-6^. A breath test for these biomarkers could provide a new tool for early detection of breast cancer that is accurate, cost-effective, and safe. There is a clinical need for new tools to detect breast cancer detection because a woman in the United States has a 1 in 8 chance of developing breast cancer during her lifetime, and early detection can improve her prospects of survival.

Carcinoma-associated fibroblasts (CAFs) in breast cancer stromal tissue have been proposed as a feasible biological source of breath biomarker VOCs (Figure 1). Bone marrow-derived mesenchymal stem cells migrate to tumor stromal tissue, where they form carcinoma-associated fibroblasts (CAFs) that stimulate aggressive carcinoma phenotypes and drive metastasis^7,8^. CAFs also cause oxidative stress by generating reactive oxygen species that oxidize polyunsaturated fatty acids (PUFAs) in cell membranes^9^. The metabolic products of PUFA oxidation include n-alkanes such as pentane and hexane, as well as alkane derivatives; all are exhaled in the breath as VOCs^10-13^.

**Figure 1.**
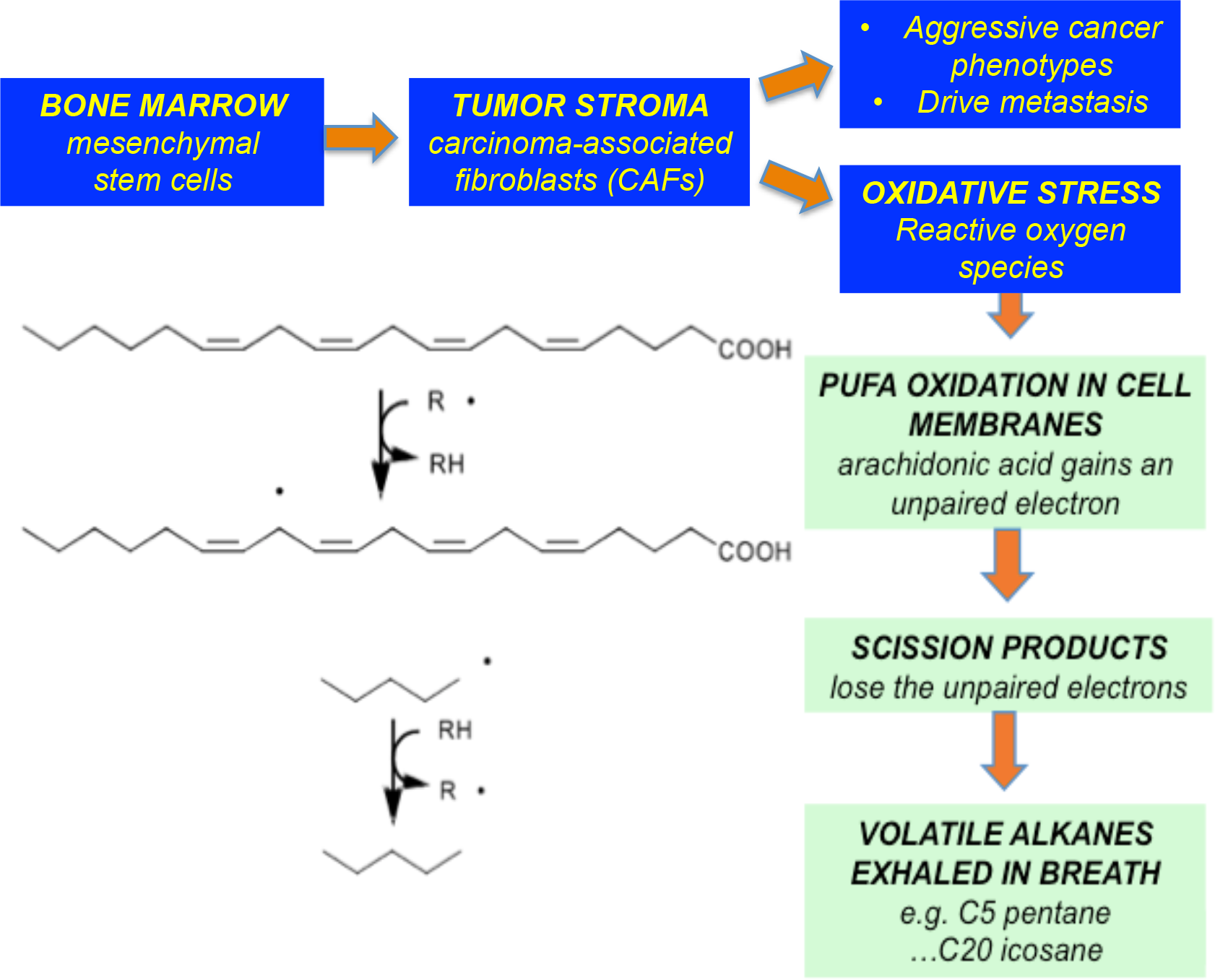
Proposed source of breath biomarkers in breast cancer: Bone marrow-derived mesenchymal stem cells migrate to tumor stromal tissue, where they form carcinoma-associated fibroblasts (CAFs) that stimulate aggressive carcinoma phenotypes and drive metastasis. CAFs also generate hydrogen peroxide and reactive oxygen species, resulting in oxidative stress. Oxidation of polyunsaturated fatty acids (PUFAs) in cell membranes generates downstream scission products include volatile n-alkanes (e.g. pentane, hexane) and their metabolic derivatives, all of which have a high vapor pressure and are excreted in the breath.

We have previously reported a rapid point-of-care breath test that detected biomarkers of breast cancer, abnormal mammograms, and pulmonary tuberculosis^4,14^. We report here a study to validate the accuracy of that test as an indicator of risk of breast cancer and abnormal mammograms in symptomatic women.

## METHODS AND MATERIALS

### Human subjects (Table 1)

We performed breath tests in 593 women with symptomatic breast disease at three sites: University of Southern California, Los Angeles, CA, MD Anderson Cancer Center, Houston, TX, and St. Michael’s Medical Center, Newark, NJ). An Institutional Review Board approved the research at all sites. A physician explained the study to women who fulfilled the inclusion and exclusion criteria and invited them to participate in the research.

**Table 1:**
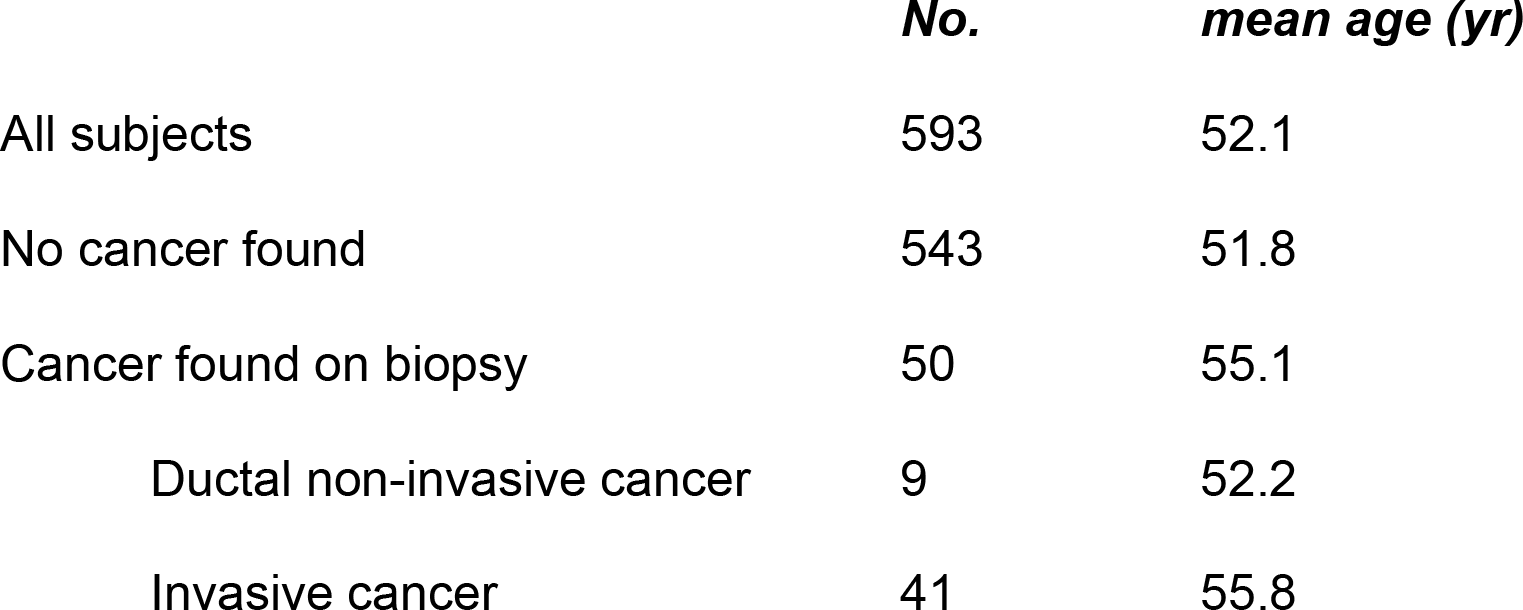
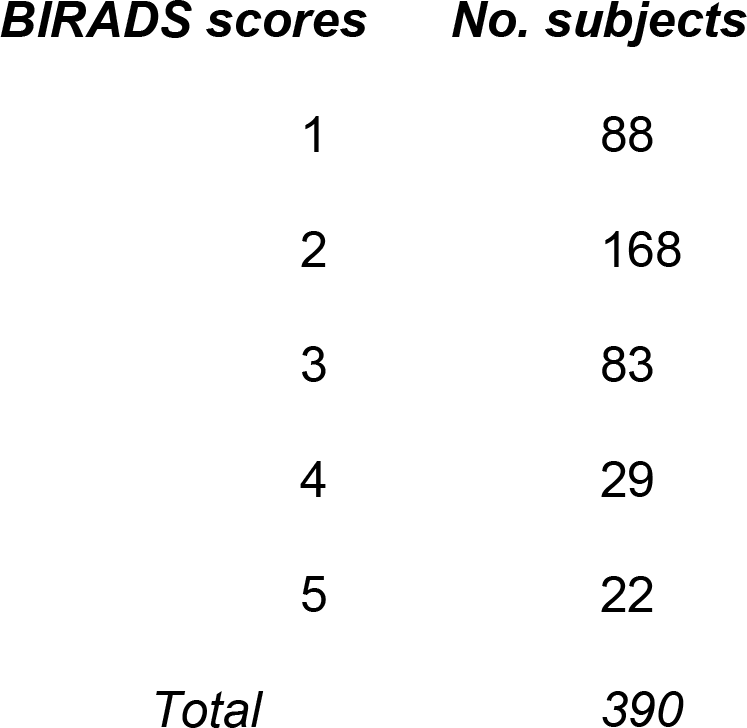
Human subjects. Breast biopsy was performed in 137 subjects, and 87 were negative for cancer. Women were classified as cancer-free if BIRADS score ≤ 3 or if the breast biopsy findings were negative. There was no significant difference between the ages of women with and without cancer (2-tailed t-test, two sample equal variance). BIRADS scores are shown for subjects who had mammography performed with radiological imaging; subjects assessed with other modalities were not included in this table.

### Inclusion criteria

Women were included in the study if they were aged 18 years or over, and had been referred for mammography for a breast-related symptom or clinical sign (e.g. a breast mass or a nipple discharge). All gave their written informed consent to participate and they approved the collection of clinically relevant data including mammogram and biopsy results.

### Exclusion criteria

Women were excluded from the study if they had a known serious or potentially life-threatening disease, a previous history of cancer (with the exception of basal cell carcinoma of skin), or if there was a history of a mammogram during the preceding 12 months.

### Breath VOC collection and analysis

The method has been described ^4,14^. Breath samples were collected and analyzed with a rapid point-of-care instrument employing gas chromatography and surface acoustic wave detection (GC SAW) (BreathLink, Menssana Research, Inc, Fort Lee, NJ) (Figure 2). The instrument automatically collected and concentrated alveolar breath VOCs onto a sorbent trap containing Tenax®, and thermally desorbed them for analysis with GC SAW in < 6 min. Breath VOC chromatograms were uploaded electronically to a central server for analysis of data. The analyzer was re-calibrated daily with an external standard, a mixture of C6 to C22 n-alkanes (Restek Corporation, Bellefonte, PA 16823, USA).

**Figure 2.**
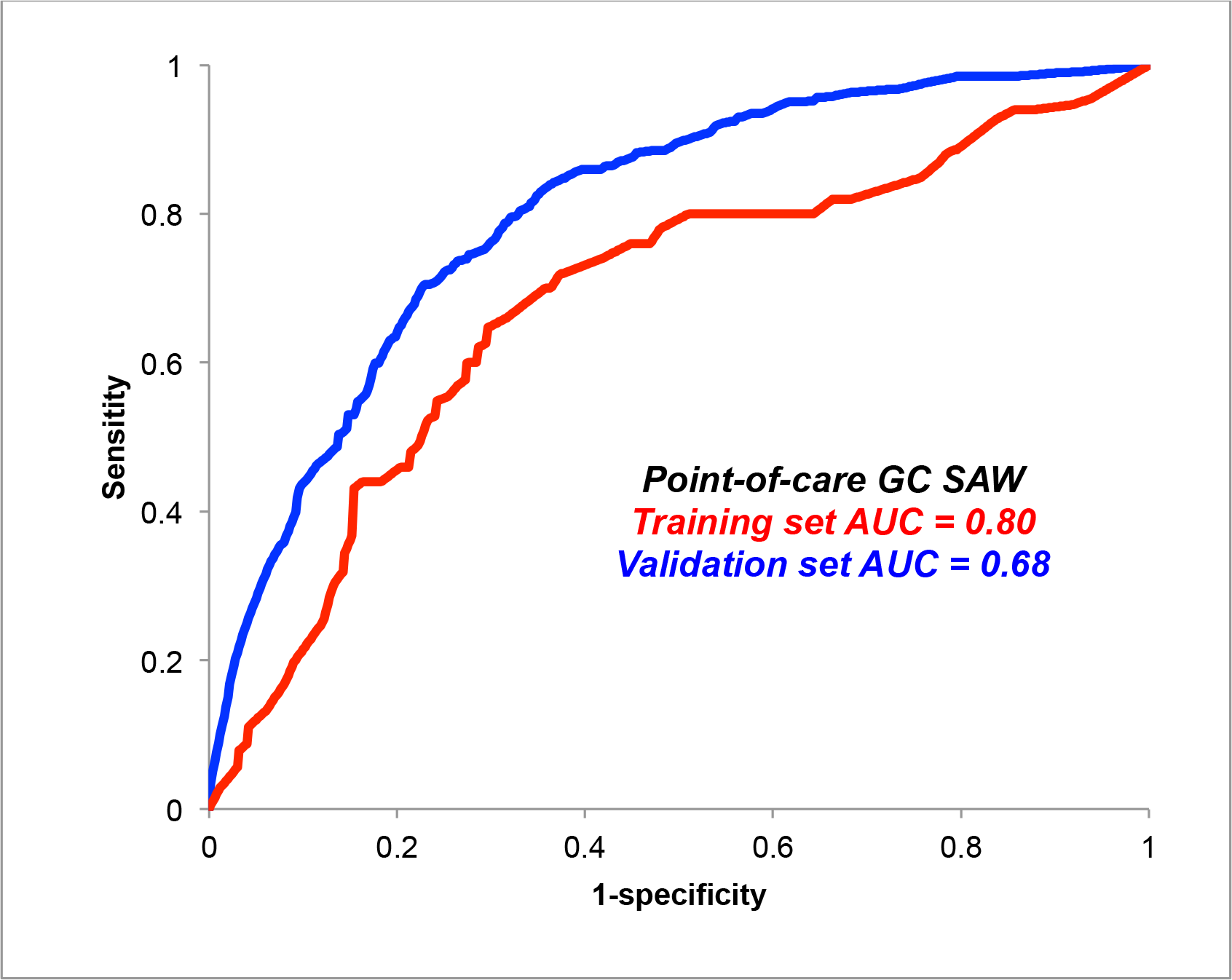
Prediction of breast cancer with breath biomarkers. This figure displays ROC curves of the sensitivity and specificity of the breath test for breast cancer, using a rapid point-of-care GC SAW test for breast cancer (BreathLink). Data was split into training and validation sets. The area under curve (AUC) of the ROC curve indicates that the breath test identified breast cancer with 83% accuracy in the training set and 68% accuracy in the validation. set

### Analysis of data

Data were analyzed to determine the accuracy of the breath test as a predictor of biopsy-proven breast cancer and also as a predictor of mammogram results. The methods have been described^4,5^. Breath VOC chromatograms were randomly assigned to a training set or a validation set. Monte Carlo analysis identified significant breath VOC biomarkers of breast cancer and abnormal mammograms in the training set, and these biomarkers were incorporated into a multivariate algorithm to predict disease in the validation set.

### Training sets

Predictive models were trained using multiple Monte Carlo simulations and multivariate weighted digital analysis (WDA)^15^. Chromatograms were aligned and binned into a time series of data segments derived from the SAW detector signal and the diagnostic accuracy of each data segment was ranked according to the area under curve (AUC) of its receiver operating characteristic (ROC). If a data segment identified disease (breast cancer or an abnormal mammogram) with greater than random accuracy (p<0.05), it was entered into a WDA multivariate predictive algorithm.

### Validation sets^4^

*Breast cancer data*. The WDA model was validated with five random 80/20 splits of the dataset and the results were averaged. *Mammography data:* Data were stratified in two ways: BIRADS 1 and 2 versus BIRADS 3, 4 and 5, and BIRADS 1, 2 and 3 versus BIRADS 4 and 5. WDA models were validated with 10-fold cross validation. Chromatograms from each group were partitioned randomly into 10 “folds” i.e. in 10 trials in which the predictive models were trained on 9 folds and validated on the remaining fold.

## RESULTS

### Prediction of breast cancer (Figure 2)

50 women had biopsy-proven breast cancer. *Unsplit data set:* Breath VOCs identified breast cancer with 83% accuracy (area under curve of receiver operating characteristic), 82% sensitivity and 77.1% specificity. *Cross-validated data:* The training set breath VOCs identified breast cancer with 80.3% accuracy, 84% sensitivity and 74.3% specificity. Corresponding values in the validation set were 68.2%% accuracy, 72.4% sensitivity and 61.5% specificity.

### Prediction of mammogram results (Figure 3)

*Prediction of BIRADS 4 and 5 mammograms (versus BIRADS 1, 2 and 3): Unsplit data set:* Breath VOCs identified abnormal mammograms with 76.2% accuracy. *Split data sets:* Breath VOCs identified abnormal mammograms with 74.2% accuracy, 73.3% sensitivity and 60% specificity. Corresponding values in the validation set were 60.5% accuracy, 64.2% sensitivity and 51% specificity.

**Figure 3.**
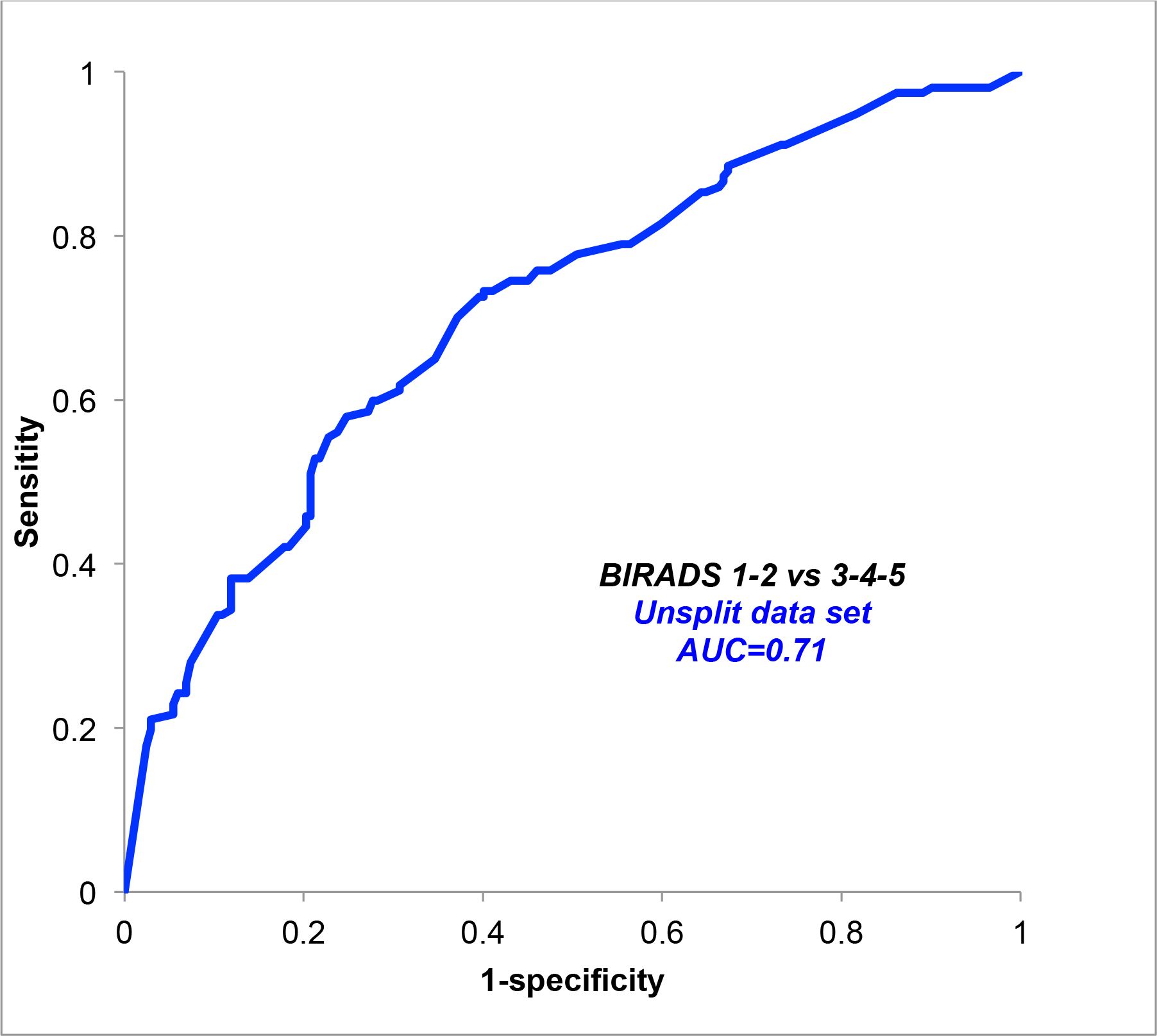
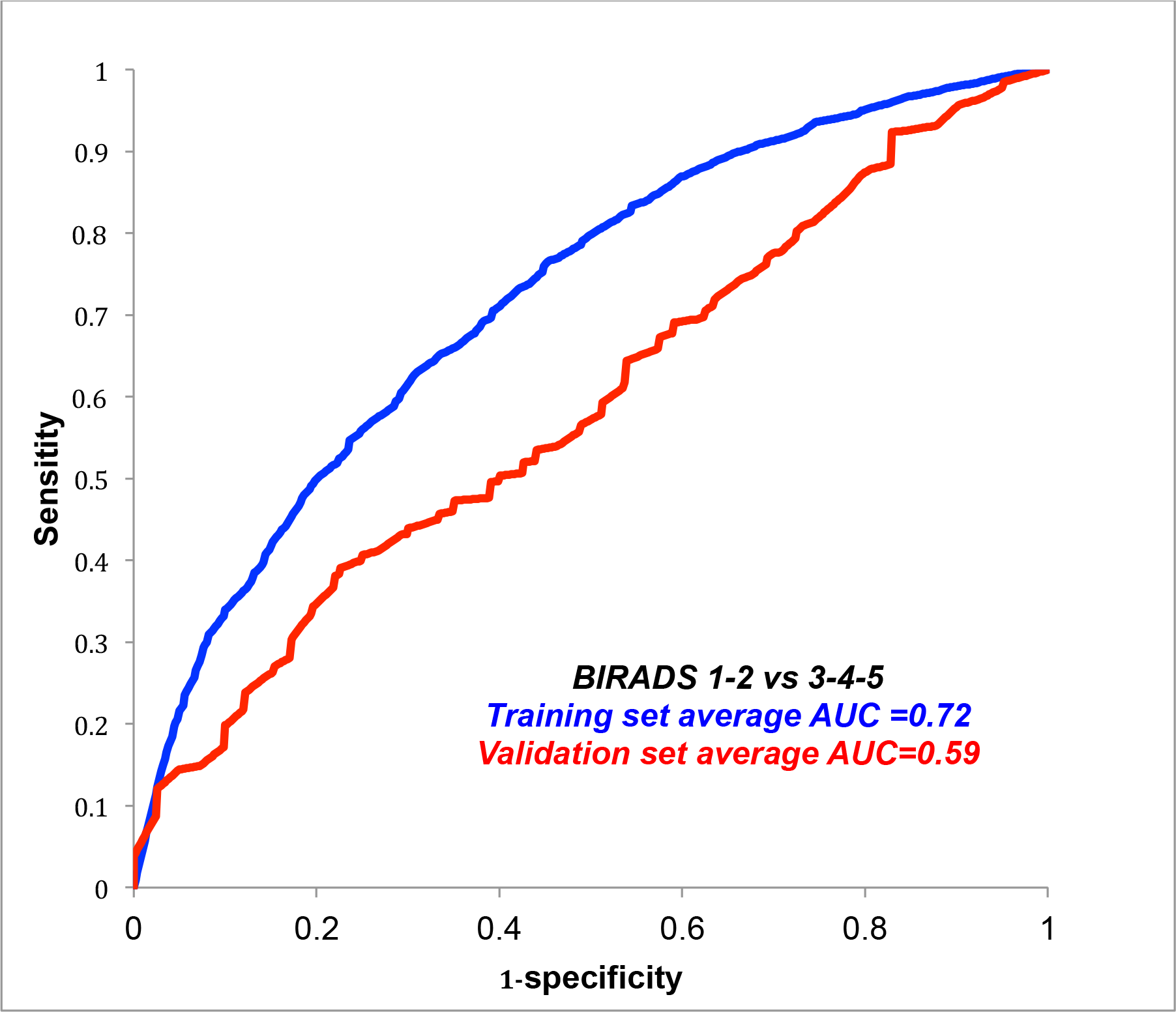
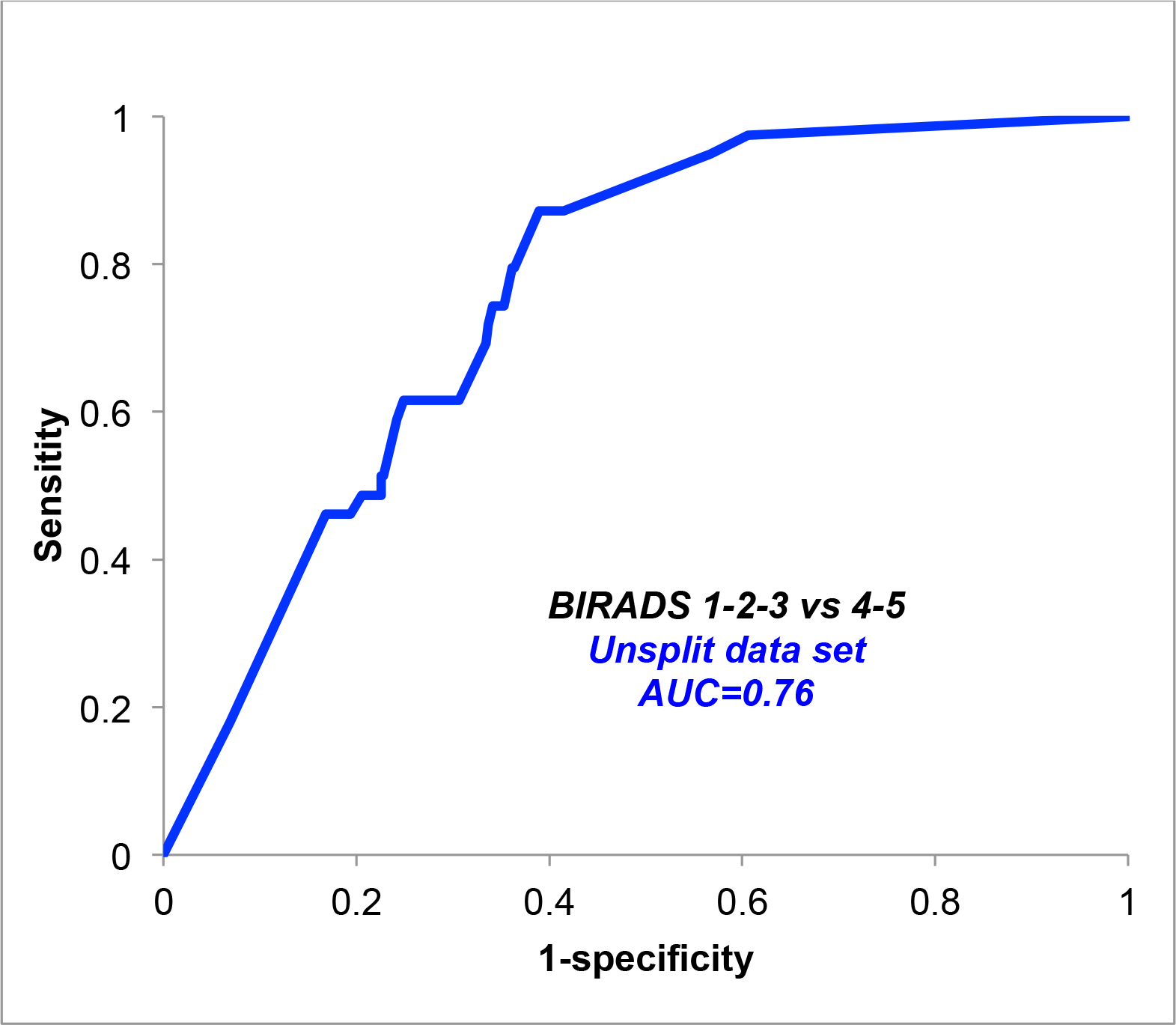
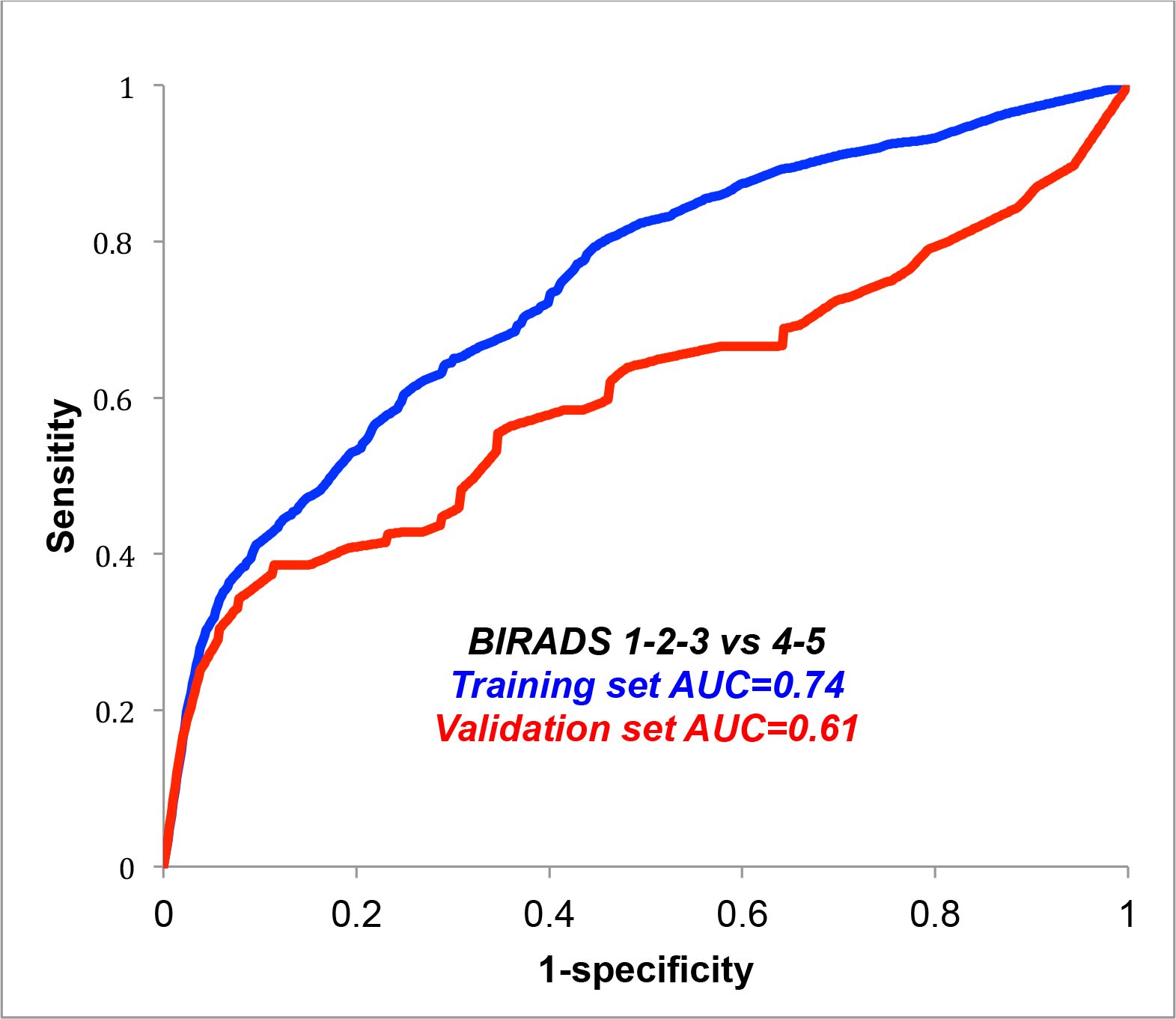
Prediction of BIRADS score with breath biomarkers. This figure displays ROC curves of the sensitivity and specificity of the breath test as a predictor of the outcome of mammography. Outcomes of the breath test were stratified in two ways: BIRADS 4 and 5 mammograms versus BIRADS 1, 2 and 3 (top left and top right panels), and BIRADS 3, 4 and 5 mammograms versus BIRADS 1 and 2 (bottom left and bottom right panels). BIRADS scores are shown for subjects who had mammography performed with radiological imaging; subjects assessed with other modalities were not included in this analysis.

*Prediction of BIRADS 3, 4 and 5 mammograms (versus BIRADS 1 and 2): Unsplit data set: Overall accuracy was 71%. Cross-validated data:* The training set and the validation set identified abnormal mammograms with 72% and 59% accuracy respectively.

### Effects of age (Table 1)

There was no significant difference between the mean ages of women with and without breast cancer.

## DISCUSSION

The main finding of this study was that a rapid point-of-care breath test for VOC biomarkers accurately predicted the risk of breast cancer and abnormal mammograms in women with breast-related symptoms. The accuracy of the breath test was consistent with values reported in previous studies.

Hietanen et al reported increased breath pentane in breast cancer in 1994^16^, and we subsequently confirmed increased n-alkanes in breath (nonane, tridecane) and methylated derivatives of n-alkanes (5-methyl undecane, 3-methyl pentadecane) as candidate biomarkers of breast cancer using GC MS^1^. Other investigators have also reported distinctive breath VOCs in breast cancer using GC MS^17 18^ as well as with nanosensor arrays^19^ and sniffing dogs^20^. The biological source of breath VOC biomarkers in breast cancer may reside in activated breast stromal fibroblasts where increased oxidative stress generates volatile n-alkanes including ethane and pentane and other metabolic products that are expired in the breath (Figure 1) ^21 11,13^. Also, headspace analysis of VOCs derived from breast cancer cells cultured in vitro has demonstrated a variety of unique products, some of which may have arisen from induced cytochrome p450 activity^22^.

The experimental design incorporated precautions that were targeted to minimize the effects of potential confounding variables. Breath tests were performed in a blinded fashion without knowledge of the results of breast biopsy. We minimized the potential effects of site-dependent confounders (e.g. ambient room air contamination) by collecting and analyzing breath samples from subjects with and without cancer in the same room at each site. We also incorporated precautions in the analysis of data by employing multiple Monte Carlo simulations to minimize the risk of “over-fitting” data when large numbers of candidate biomarkers are correlated with a comparatively small number of experimental subjects. In the absence of this precaution, there is a risk of generating “voodoo correlations” in which the findings appear to be statistically significant even though they are clinically meaningless^23^. In addition, we cross-validated the test results with multiple random splits of the data sets into training sets and validation sets in order to ensure that the predictive algorithms were developed and tested in independent groups of subjects. There was no significant difference between the mean ages of women with and without breast cancer

In this study, a rapid point-of-care test for breath VOC biomarkers was a sensitive and specific predictor of risk of breast cancer and abnormal mammograms in women with breast-related symptoms. These findings support the use of breath VOC biomarker analysis as a tool for stratifying a screening population into groups at low or high risk of breast cancer. Breath testing is painless, cost-effective, and completely safe, and it could potentially reduce the number of needless mammograms that are now performed.

## Data Availability

Non-proprietary data is available from Menssana Research Inc.

## Compliance with Ethical Standards

### Funding

This study was funded by NIH NCI Grant Number: 5R44CA203019 – 02

### Conflict of Interest

Michael Phillips is President and CEO of Menssana Research, Inc. All other authors declare that they have no conflict of interest.

### Animals

No animals were involved.

### Human subjects

The study was reviewed and approved by an Institutional Review Board (IRB) at all participating sites. Written informed consent was obtained from all individual participants included in the study. All procedures performed in studies involving human participants were in accordance with the ethical standards of the institutional and/or national research committee and with the 1964 Helsinki declaration and its later amendments or comparable ethical standards.

## Acknowledgements

Michael Phillips is President and CEO of Menssana Research, Inc. Schmitt & Associates, Newark, NJ, maintained a database of chromatograms and Daniel Strano and Jonah Phillips analyzed the data. Funding source: NIH NCI Grant Number: 5R44CA203019 – 02. ClinicalTrials.gov Identifier: NCT02888366

## Ethics and reporting

All relevant ethical guidelines were followed, all necessary IRB approvals were obtained, all necessary patient consent was been obtained and the appropriate institutional forms were archived.

## Notes

### Competing Interest Statement

Michael Phillips is CEO of Menssana Research Inc.

### Clinical Trial

NCT02888366

## REFERENCES

1. Phillips M, Cataneo R, Ditkoff B, et al. Volatile markers of breast cancer in the breath. Breast J 2003;9:184–91.

2. Phillips M, Cataneo RN, Ditkoff BA, et al. Prediction of breast cancer using volatile biomarkers in the breath. Breast Cancer Res Treat 2006;99:19–21.

3. Phillips M, Cataneo RN, Saunders C, Hope P, Schmitt P, Wai J. Volatile biomarkers in the breath of women with breast cancer. J Breath Res 2010;4:026003.

4. Phillips M, Beatty JD, Cataneo RN, et al. Rapid point-of-care breath test for biomarkers of breast cancer and abnormal mammograms. PLoS One 2014;9:e90226.

5. Phillips M, Cataneo R, Lebauer C, Mundada M, Saunders C. Breath mass ion biomarkers of breast cancer. J Breath Res 2016 Dec 19 doi: 101088/1752-7163/aa549b [Epub ahead of print] 2016.

6. Peng G, Hakim M, Broza YY, et al. Detection of lung, breast, colorectal, and prostate cancers from exhaled breath using a single array of nanosensors. Br J Cancer 2010;103:542–51.

7. Barcellos-de-Souza P, Comito G, Pons-Segura C, et al. Mesenchymal Stem Cells are Recruited and Activated into Carcinoma-Associated Fibroblasts by Prostate Cancer Microenvironment-Derived TGF-beta1. Stem Cells 2016;34:2536–47.

8. Sasaki T, Franco OE, Hayward SW. Interaction of prostate carcinoma-associated fibroblasts with human epithelial cell lines in vivo. Differentiation 2017;96:40–8.

9. Du C, Guo Y, Cheng Y, Han M, Zhang W, Qian H. Torulene and torularhodin, protects human prostate stromal cells from hydrogen peroxide-induced oxidative stress damage through the regulation of Bcl-2/Bax mediated apoptosis. Free Radic Res 2017;51:113–23.

10. Kneepkens CM, Ferreira C, Lepage G, Roy CC. The hydrocarbon breath test in the study of lipid peroxidation: principles and practice. Clin Invest Med 1992;15:163–86.

11. Kneepkens CM, Lepage G, Roy CC. The potential of the hydrocarbon breath test as a measure of lipid peroxidation. Free Radic Biol Med 1994;17:127–60.

12. Chan HP, Lewis C, Thomas PS. Oxidative stress and exhaled breath analysis: a promising tool for detection of lung cancer. Cancers (Basel) 2010;2:32–42.

13. Aghdassi E, Allard JP. Breath alkanes as a marker of oxidative stress in different clinical conditions. Free Radic Biol Med 2000;28:880–6.

14. Phillips M,., Basa-Dalay V, Blais J, et al. Point-of-care breath test for biomarkers of active pulmonary tuberculosis. Tuberculosis (Edinburgh) 2012;92:314–20.

15. Phillips M, Altorki N, Austin JH, et al. Detection of lung cancer using weighted digital analysis of breath biomarkers. Clin Chim Acta 2008;393:76–84.

16. Hietanen E, Bartsch H, Bereziat JC, et al. Diet and oxidative stress in breast, colon and prostate cancer patients: a case-control study. Eur J Clin Nutr 1994;48:575–86.

17. Patterson SG, Bayer CW, Hendry RJ, et al. Breath analysis by mass spectrometry: a new tool for breast cancer detection? Am Surg 2011;77:747–51.

18. Mangler M, Freitag C, Lanowska M, Staeck O, Schneider A, Speiser D. Volatile organic compounds (VOCs) in exhaled breath of patients with breast cancer in a clinical setting. Ginekol Pol 2012;83:730–6.

19. Xu Y, Lee H, Hu Y, Huang J, Kim S, Yun M. Detection and identification of breast cancer volatile organic compounds biomarkers using highly-sensitive single nanowire array on a chip. J Biomed Nanotechnol 2013;9:1164–72.

20. McCulloch M, Jezierski T, Broffman M, Hubbard A, Turner K, Janecki T. Diagnostic accuracy of canine scent detection in early- and late-stage lung and breast cancers. Integr Cancer Ther 2006;5:30–9.

21. Jezierska-Drutel A, Rosenzweig SA, Neumann CA. Role of oxidative stress and the microenvironment in breast cancer development and progression. Adv Cancer Res 2013;119:107–25.

22. Silva CL, Perestrelo R, Silva P, Tomas H, Camara JS. Volatile metabolomic signature of human breast cancer cell lines. Scientific reports 2017;7:43969.

23. Miekisch W, Herbig J, Schubert JK. Data interpretation in breath biomarker research: pitfalls and directions. J Breath Res 2012;6:036007.

